# Clinical Methods Supporting Recognition of Early Post-Stroke Seizures: A Systematic Scoping Review

**DOI:** 10.1101/2023.04.25.23289090

**Authors:** Clare Gordon, Hedley Emsley, Catherine Elizabeth Lightbody, Andrew Clegg, Catherine Harris, Joanna Harrison, Catherine Emma Davidson, Jasmine Wall, Caroline Watkins

## Abstract

**Background:** Stroke is one of the commonest cause of seizures and epilepsy and is the leading cause of epilepsy over the age of 60 (1, 2). Post-stroke seizures and epilepsy are associated with increased mortality, disability and recurrent hospital admissions (3, 4). Seizures occurring in the immediate aftermath of the acute stroke can complicate a patient’s stroke diagnosis and management or can even go undiagnosed resulting in increased risk of mortality, disability and hospital readmissions. There is limited evidence on detection, observation, diagnosis and management of early post-stroke seizures as part of acute stroke treatment.

**Objectives:** The objective of this series of scoping reviews is to map the extent and type of literature in relation to in-hospital early post-stroke seizures. For this paper, the specific objectives relate to the clinical methods used in the bedside identification and observation, usually performed by nurses, of early post-stroke seizures (EPSS) in adults being treated and managed for acute stroke.

**Eligibility criteria:** Participants included adults aged 18 years or older with acute ischaemic stroke or primary intracerebral haemorrhage and a diagnosis, or suspected diagnosis, of post-stroke seizures whilst receiving hospital care for their acute stroke.

**Sources of evidence:** Medline, CINAHL, Embase, and the Cochrane Library databases were searched, including papers published up to October 2021, limited to English language. A broad range of published literature was selected comprising of primary research, including case studies/case reports, conference abstracts, systematic reviews/meta-analyses, clinical guidelines and consensus statements. Reference lists of included studies were also searched.

**Charting methods:** A data charting table was developed by the reviewers, with key information selected for included articles. Findings have been aggregated to an overview of extent and type of evidence and identify gaps in evidence.

**Results:** We included two research papers, two clinical guidelines and four discussion papers. There was limited literature on clinical methods used to identify and observe acute stroke patients for seizures. We found no evaluation of different methods aimed at recognising and observing EPSS, and subsequently recommendations lacking detail and consensus on clinical processes.

**Conclusion:** Early post-stroke seizures are important to diagnose due to associated increases in post-stroke complications, mortality, disability and recurrent hospital admissions. Whilst the diagnostic challenge of EPSS is recognised, there is a need for research looking into how to improve the identification and observation of seizure activity in acute stroke settings.

## INTRODUCTION

Early post-stroke seizures (EPSS), also termed provoked or acute symptomatic seizures, describe seizures provoked by the acute brain injury associated with a stroke, whereas post-stroke epilepsy arises from long-term changes to the brain after stroke with recurrent seizures that are unprovoked by any other factor, such as metabolic, toxic etc. (5, 6). Post-stroke seizures occurring in the immediate aftermath of the acute stroke can complicate a patient’s stroke diagnosis and management or can even go undiagnosed. They may cause new or seemingly unexplained persistence of focal deficits (due to post-ictal paresis), or reduced consciousness, which can be complicated to assess and distinguish from the neurological deficit of the stroke itself leading to diagnostic challenge. There is a lack of consensus in the literature on the definition and timing of what is classed as EPSS or epilepsy, with EPSS defined as occurring between 48 hours and two weeks after acute stroke (5, 7). Whereas the International League Against Epilepsy defines early post-stroke seizures as up to seven days after stroke onset (8).

The risk of developing post-stroke epilepsy is substantially higher in patients presenting with an early seizure than in patients with stroke and no early seizure (6). EPSS are more likely to occur after intracerebral haemorrhage (prevalence 10-16% across stroke populations) but are also common after ischaemic stroke (prevalence 3-15%), in ischaemic stroke with haemorrhagic transformation, cortical involvement and with increasing stroke severity (6, 9). Numerically, given the epidemiology of pathological stroke subtypes, early post-ischaemic stroke seizures will be more frequently encountered in the acute stroke context. It is important to identify and diagnose EPSS as they are associated with increased mortality, disability and recurrent hospital admissions (3, 10). Accurately determining the prevalence of post-stroke seizures, and effective treatment of seizures to prevent their associated complications, depends on the methods used to identify and diagnose seizures and guidance on an agreed systematic approach for clinical practice is currently lacking (5, 11, 12).

In the context of the absence of an agreed definition for EPSS and lack of reference specifically to EPSS in national guidance, this study was undertaken as part of a series of three reviews, which aimed to systematically scope the practice and research literature on in-hospital EPSS, to identify current knowledge on its clinical recognition and diagnosis, map the inclusion of EPSS within national and international clinical guidelines, and identify current evidence on its in-hospital management.

Our focus for this sub-study was to map the breadth of evidence in relation to clinical methods used to support identification and observation of early post-stroke seizures (EPSS) and identify gaps in this evidence. Our research question was: *What is known from the existing literature about the clinical methods used for identifying and observing seizures in adults being treated for acute stroke?*

## METHODS

The review was guided by Arksey and O’Malley’s framework which includes: (i) identifying the research question; (ii) searching for relevant papers; (iii) selecting papers; (iv) charting the data; (v) collating, summarising and reporting the results (13, 14). The inclusion criteria and methods were pre-specified and published online (OSF ID: bkejc) (15).

### Identifying the research question and eligibility criteria

The research question was developed with input from subject experts comprising of academics and clinicians and patient and public involvement (PPI) for the identification of relevant outcomes. We included practice (i.e., clinical guidelines) and research literature involving adults (>18 years) with acute stroke (ischaemic or primary intracerebral haemorrhage) and seizures that occurred in hospital, within two weeks of stroke onset. We included seizures occurring at stroke onset and seizures occurring with an acute stroke intervention such as reperfusion therapies. We excluded literature reporting on seizure as a stroke mimic, patients with known epilepsy or seizures before their stroke, and patients with diagnosis of subdural and subarachnoid haemorrhages or cerebral ischaemia without arterial circulation obstruction, such as vasospasm or secondary to trauma. We included literature published up to October 2021, all study types including systematic reviews with meta-analysis and non-research literature such as clinical guidelines and consensus statements. We excluded papers published in languages other than English. See Table 1 for key elements of the review question.

**Table 1.** PICo criteria for the research aim.

| Population | Interest | Context |
| --- | --- | --- |
| Adults ($\geq 18$ yrs)<br>Acute ischaemic stroke<br>Primary intracerebral haemorrhage (haemorrhagic stroke)<br>Diagnosis / suspected early post-stroke seizures or epilepsy | Prevalence<br>Incidence<br>Seizure recognition/ identification<br>Seizure/epilepsy diagnosis<br>Management of early seizures/epilepsy | Receiving in-patient hospital care for acute stroke<br>Less than 2 weeks post stroke onset |

### Searching for relevant papers

MEDLINE (Ovid), CINAHL (EBSCOhost), EMBASE (Ovid) and the Cochrane Library databases were searched up to 1^st^ October 2021. To ensure that all relevant information was captured, we also searched a variety of grey literature sources (searched January 2022): Grey Literature Report, OpenGrey and Web of Science Conference Proceedings to identify studies, case reports and conference abstracts of relevance to this review. We also conducted a targeted search using Google of the grey literature and specifically searched national and international organisations’ websites with an interest in stroke and/or seizures, such as the Stroke Association, the Epilepsy Society, the International League Against Epilepsy, the British and Irish Association of Stroke Physicians, the European Stroke Organisation and the American Stroke Association. A hand search was conducted using the reference lists of included papers to identify additional relevant papers. The search strategy was developed and piloted by an information specialist (CH) with input from the project team. The search strategies are provided in Appendix 2 and are published online (15).

### Selection of sources of evidence and charting the data

Following the searches, duplicate records were removed in EndNote before results were uploaded into Rayyan© online collaborative systematic review software (16) for record management and title and abstract screening. A two-part screening process against the inclusion criteria was used: (a) a title and abstract review and (b) full text review.

Title and abstract screening were conducted mainly by one reviewer, with 1000 citations independently screened by two reviewers (CG & JW) with 91.5% agreement between reviewers. Full text papers were assessed against the inclusion criteria, reasons for exclusion were recorded and are reported in the results by one reviewer. Any disagreements that arose were resolved through discussion with the wider project team. The results of the search selection are reported using the Preferred Reporting Items for Systematic Reviews and Meta-analyses extension for scoping review (PRISMA-ScR) flow diagram (17). The completed PRISMA-ScR checklist is presented in Appendix 1.

### Data charting process

We developed and piloted our data charting form with evidence synthesis experts (JH & AC) based on scoping review methodology (13). The piloting process included data charting of three papers independently with comparison of accuracy and comprehension after completion. Charting of the data was divided between the three reviewers (CG, JW, CD). Where results of the same study were reported in more than one publication, we collated the results and used the publication with the most data relevant to our research question as the primary reference. Data charted included type of paper (e.g., primary research, conference proceedings, clinical guideline), study aims, methods, clinical assessment method, participants, study location, study setting, type of stroke, type of seizure and key results relevant to our research question (e.g., sensitivity and specificity of tool). A scoping review does not typically involve a quality assessment and therefore we did not appraise the quality of evidence (14). Due the large number of records screened (n=15,033), included records were grouped into the following categories 1. Clinical methods, 2. Diagnosis, 3. Management and 4.

Epidemiology. Some records were categorised into more than one category. This paper will report results on the first category - clinical methods to support identification and observation of EPSS. All data is reported in a narrative format.

## RESULTS

### Selection of sources of evidence

We included 617 papers, eight of which were categorised as papers on clinical methods used to support bedside identification and observation of seizures. The selection process is outlined in the PRISMA-SR diagram (Figure 1). Date of publication ranged from 2002-2021. Papers were from three different countries, six from the United States of America (USA) and two from Europe (Belgium, Switzerland). Four papers were discussion papers, two primary research and two clinical guidance. Three discussion papers provided recommendations on observation methods of EPSS in critical care settings and one (18) in specialist stroke services (Table 2).

**Figure 1.**
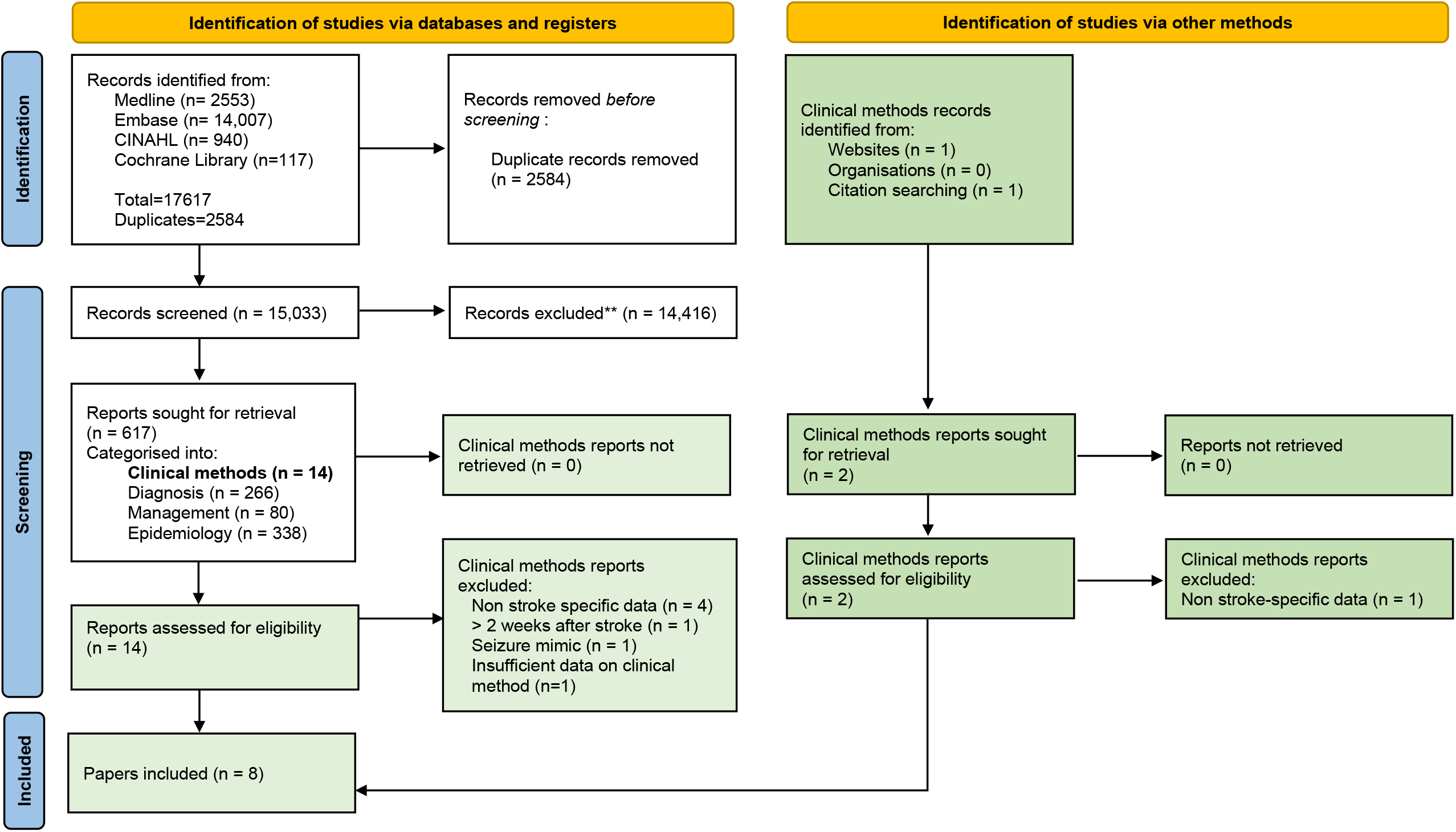
PRISMA-SR diagram to show the selection process for the review.

**Table 2.** Discussion papers for the identification and observation of early post stroke seizures.

| First author, year | Country | Clinical setting | Clinical method to identify seizure | Duration of method | Indications |
| --- | --- | --- | --- | --- | --- |
| Kraus, 2002 | USA | Critical care | Continuous EEG | Not reported | Not reported |
| Vespa, 2005 | USA | Critical care | Continuous EEG on monitor at bedside, nurse continuous review with routine periodic review by physician | 5 days | Lack of clinical seizure activity not an indication to avoid cEEG |
| De Reuck,<br>2009 | Belgium | Stroke | EEG | Not reported | As soon as possible after the ictal event |
| Bautista,<br>2020 | USA | Critical care | Nurse seizure assessment<br>Bedside EEG recording | Continuous 1<sup>st</sup> five mins and until returned to baseline | On admission |

For the two research papers, one study recruited participants from a single stroke unit, and the case study reported on a patient in the emergency department (Table 3). One clinical guideline provided guidance on acute ischaemic stroke and the second on primary intracerebral haemorrhage (19, 20). Both guidance papers were from the USA (Table 4).

**Table 3.** Studies on clinical methods for the identification and observation of early post stroke seizures.

| First author, year | Country | Study type | Study aim | Number of participants | Male (%) | Ischaemic stroke (%) | Clinical method to identify seizure |
| --- | --- | --- | --- | --- | --- | --- | --- |
| Carrera, 2006 | Switzerland | Case control | To determine the incidence and risk factors of electrical seizures and other electrical epileptic activity using continuous EEG (cEEG) in patients with acute stroke | 100 | 58 (58) | 91 (91) | Continuous EEG |
| Mader, 2020 | USA | Case report | To describe a case of stroke-onset seizures during acute ischemic stroke of the midbrain to illustrate how stroke-onset seizures might differ from other early-onset seizures | 1 | 0 (0) | 1 (100) | Spouse witness<br>Continuous EEG |

**Table 4.** Clinical guidelines for the identification and observation of early post stroke seizures.

| First author, year | Country | Clinical setting | Clinical method to identify seizure | Duration of method | Indications |
| --- | --- | --- | --- | --- | --- |
| Hemphill, 2015 | USA | Acute stroke | Continuous EEG | At least 24 hours | Depressed mental status out of proportion to the stroke |
| Green, 2021 | USA | Acute stroke | Standardised approach to recognition, assessment and documentation of the seizure<br>Neurological examination<br>EEG | Not reported | Monitor with EEG for change in mental status or depressed LoC out of proportion to the stroke |

**Table 5.**
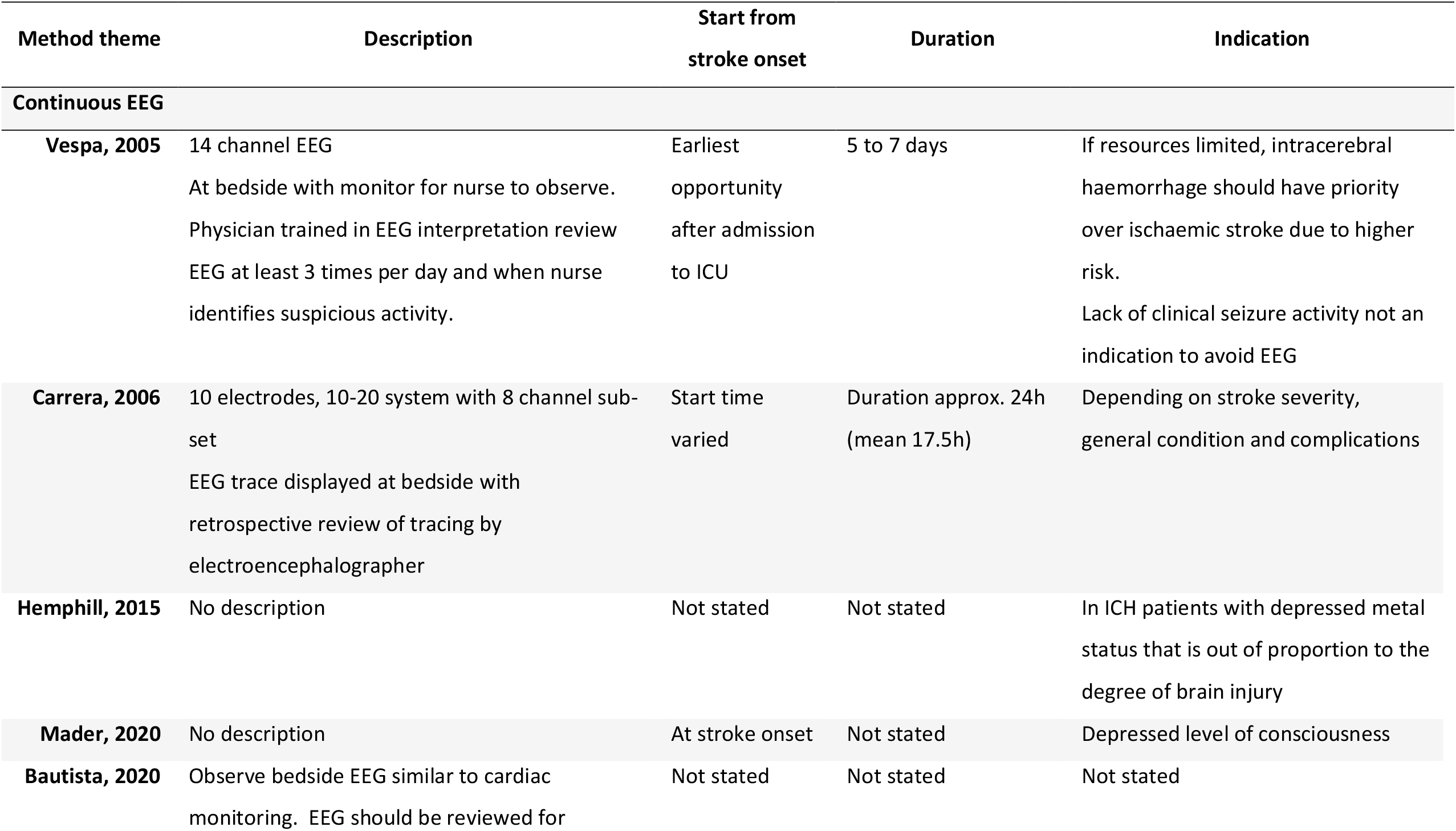
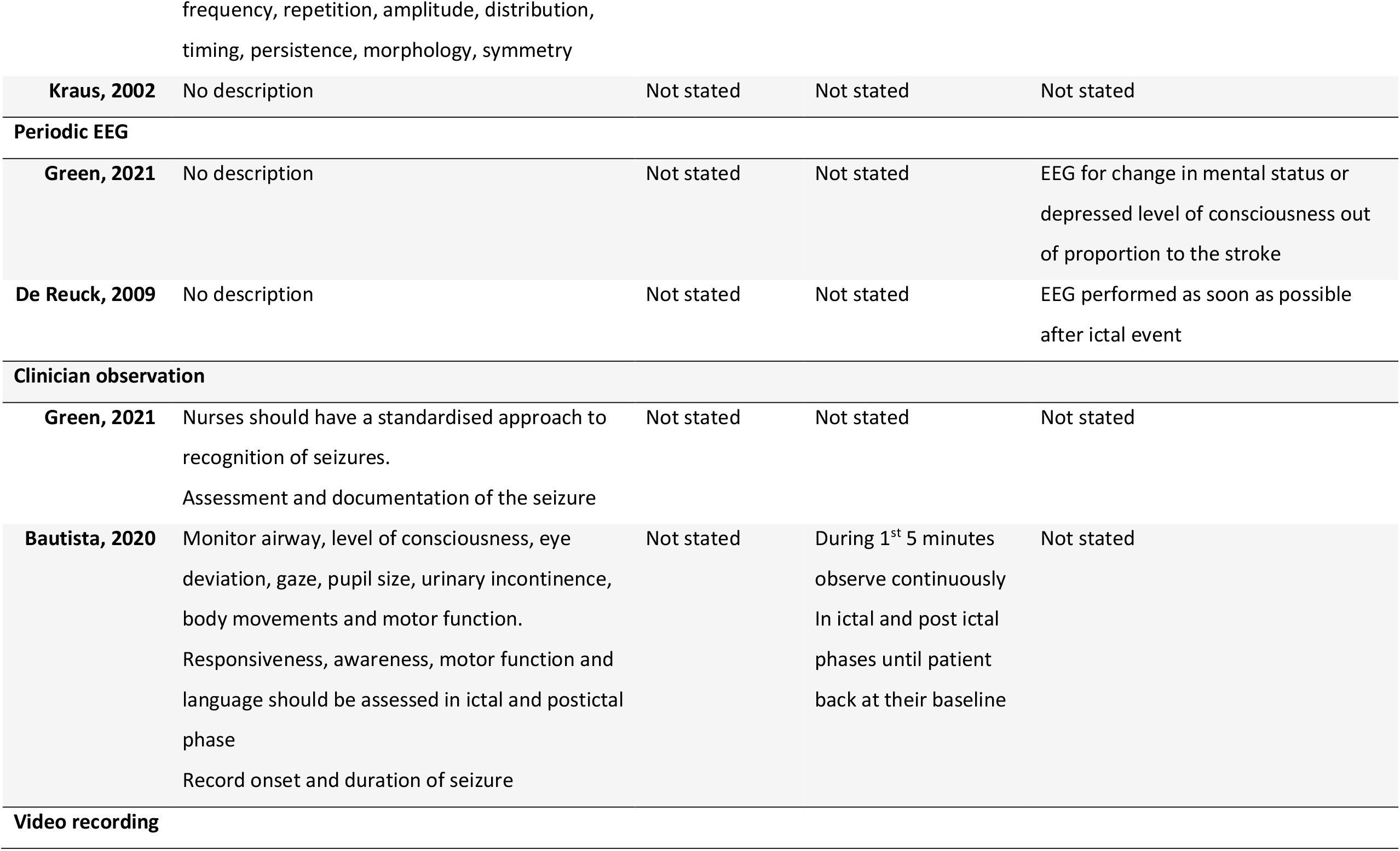
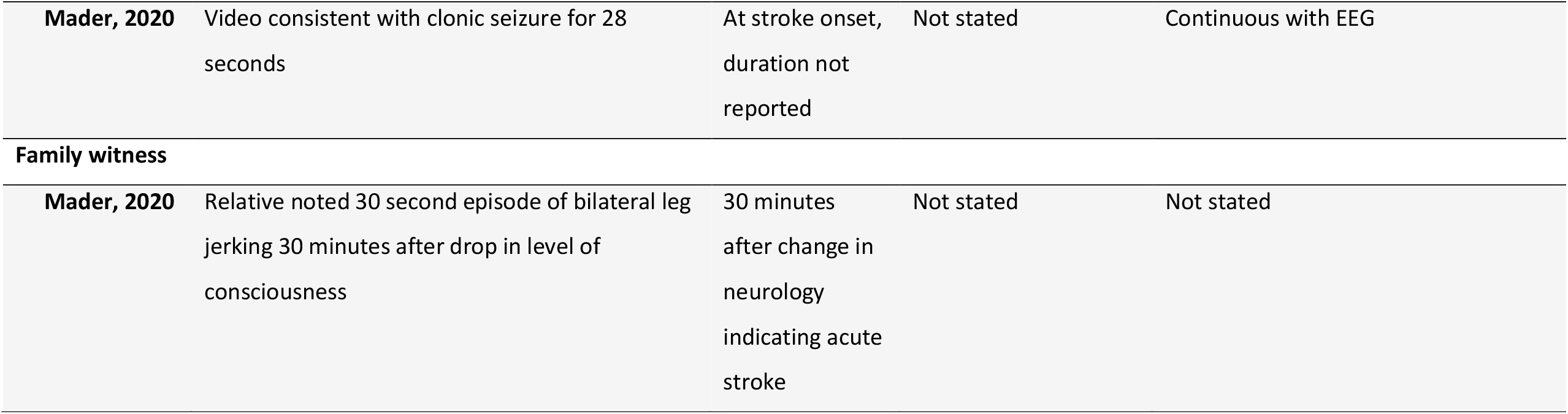
Themes of clinical methods for the identification and observation of early post stroke seizures.

| Method theme | Description | Start from stroke onset | Duration | Indication |
| --- | --- | --- | --- | --- |
| **Continuous EEG** |  |  |  |  |
| **Vespa, 2005** | 14 channel EEG<br>At bedside with monitor for nurse to observe.<br>Physician trained in EEG interpretation review EEG at least 3 times per day and when nurse identifies suspicious activity. | Earliest opportunity after admission to ICU | 5 to 7 days | If resources limited, intracerebral haemorrhage should have priority over ischaemic stroke due to higher risk.<br>Lack of clinical seizure activity not an indication to avoid EEG |
| **Carrera, 2006** | 10 electrodes, 10-20 system with 8 channel sub-set<br>EEG trace displayed at bedside with retrospective review of tracing by electroencephalographer | Start time varied | Duration approx. 24h (mean 17.5h) | Depending on stroke severity, general condition and complications |
| **Hemphill, 2015** | No description | Not stated | Not stated | In ICH patients with depressed mental status that is out of proportion to the degree of brain injury |
| **Mader, 2020** | No description | At stroke onset | Not stated | Depressed level of consciousness |
| **Bautista, 2020** | Observe bedside EEG similar to cardiac monitoring. EEG should be reviewed for | Not stated | Not stated | Not stated |
| **Kraus, 2002** | No description | Not stated | Not stated | Not stated |
| **Periodic EEG** |  |  |  |  |
| **Green, 2021** | No description | Not stated | Not stated | EEG for change in mental status or depressed level of consciousness out of proportion to the stroke |
| **De Reuck, 2009** | No description | Not stated | Not stated | EEG performed as soon as possible after ictal event |
| **Clinician observation** |  |  |  |  |
| **Green, 2021** | Nurses should have a standardised approach to recognition of seizures.<br>Assessment and documentation of the seizure | Not stated | Not stated | Not stated |
| **Bautista, 2020** | Monitor airway, level of consciousness, eye deviation, gaze, pupil size, urinary incontinence, body movements and motor function.<br>Responsiveness, awareness, motor function and language should be assessed in ictal and postictal phase<br>Record onset and duration of seizure | Not stated | During 1<sup>st</sup> 5 minutes observe continuously<br>In ictal and post ictal phases until patient back at their baseline | Not stated |
| **Video recording** |  |  |  |  |
| **Mader, 2020** | Video consistent with clonic seizure for 28 seconds | At stroke onset, duration not reported | Not stated | Continuous with EEG |
| **Family witness** |  |  |  |  |
| **Mader, 2020** | Relative noted 30 second episode of bilateral leg jerking 30 minutes after drop in level of consciousness | 30 minutes after change in neurology indicating acute stroke | Not stated | Not stated |

#### Characteristics of primary research papers

Table 3 provides a summary of the key characteristics. Neither research study had a primary aim to evaluate methods of recognising and observing seizures as part of usual nursing observations.

Participants’ ages ranged from 31-94 years and a mean age of 69. Ethnicity was not reported. The case control study (21) had 99% of the participants and had a mixed sample of ischaemic and 9% (n=9) haemorrhagic stroke. Its aim was to determine the incidence of electrical seizures and epileptic electrical activity using continuous electroencephalogram (cEEG) (21). Although the paper did not evaluate the methods used for seizure recognition and monitoring, the authors provided a narrative description of the methods used in the study. The second research paper, a case study, reported on an unusual case of stroke-onset seizure from acute midbrain infarction and provided a qualitative description of the process of seizure recognition and observation accompanied with diagnostic cEEG monitoring (22).

#### Synthesis of results on clinical methods of identification and observation of seizures

Our main aim for this review was to map available literature on methods used in the identification and bedside observation of EPSS, usually performed by nurses. The review demonstrates a significant lack of coverage in the literature in this specific area of clinical practice. No records were retrieved that evaluated the accuracy of different clinical methods. Five method types were identified: (i) continuous EEG, (ii) periodic EEG, (iii) clinician (nurse) observation, (iv) video recording, and (v) family witness. Continuous EEG was the most frequently occurring method type (Figure 2).

**Figure 2.**
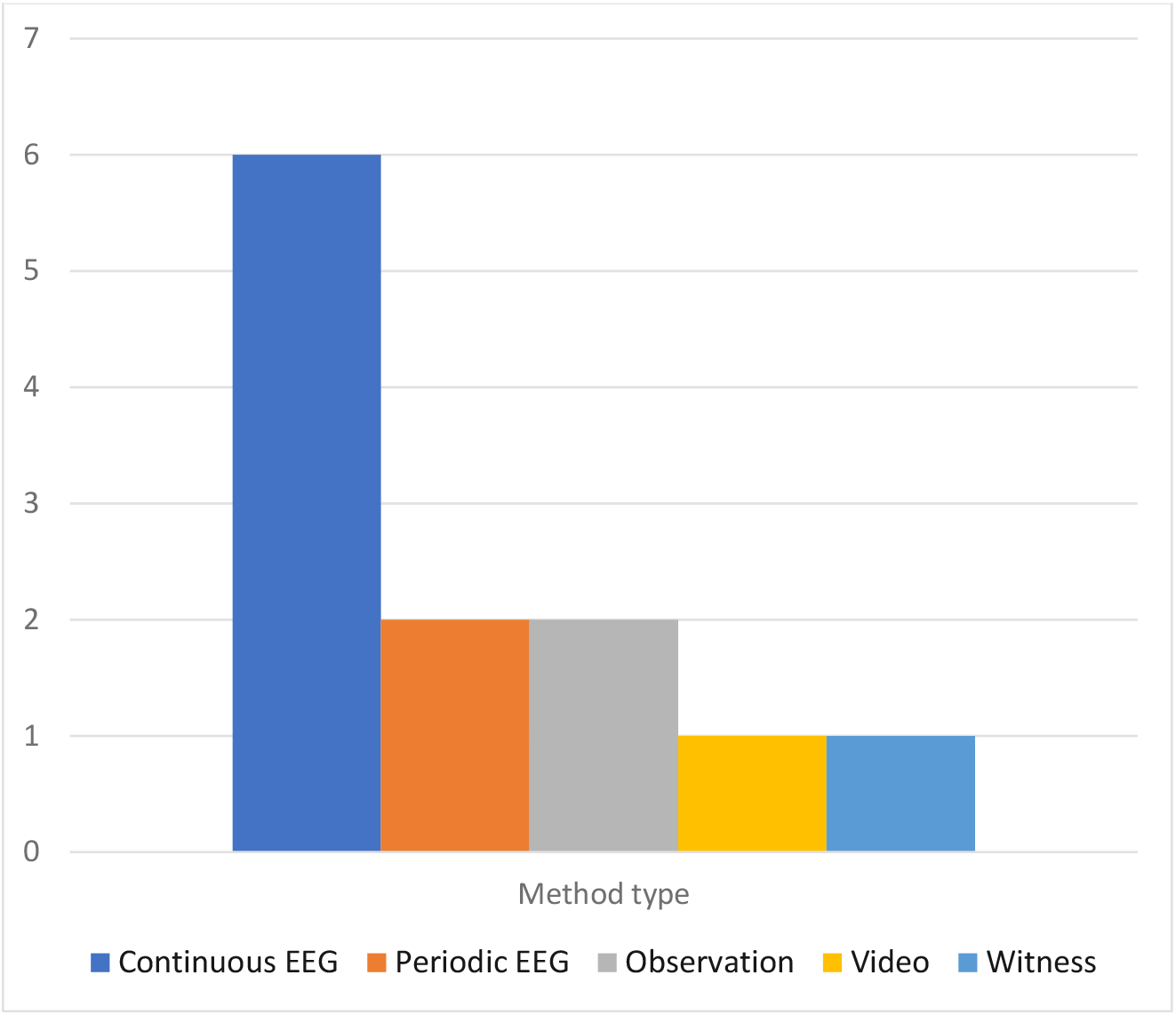
Frequency of occurrence of methods for identifying and observing seizures in the eight included papers

Table 4 provides a summary of the key information for each method.

i. Continuous EEG (cEEG). Six papers referred to cEEG to identify and observe EPSS. Five of these papers were from the USA. Indications and duration of cEEG varied; either cEEG is commenced routinely at the earliest opportunity after stroke (22, 23) or commenced depending on clinical complications, usually depressed level of consciousness (20–22). There was consensus that cEEG monitoring should be used similarly to continuous cardiac (ECG) monitoring. Type of cEEG and duration of monitoring was only reported in two papers (21, 23): cEEG ranged from 14–8 channels and duration from 24h to 7 days. Three papers described cEEG at the bedside with a monitor for nursing observation (21, 23, 24) and two described nurses requiring skills to identify electrical seizure activity (23, 24). Bautista (2020) outlined EEG knowledge required on frequency, repetition, amplitude, distribution, timing, persistence, morphology, and symmetry (24). In addition to nurse observation, two papers described retrospective review of the cEEG by either a physician trained in EEG interpretation or by an electroencephalographer (21, 23).
ii. Periodic EEG. Two papers referred to periodic EEG for either change in level of consciousness out of proportion to the stroke (Green 2021), or to be conducted as soon as possible after an EPSS (18). No details were given regarding the EEG channel system.
iii. Clinician observation. Two papers described the type of nurse clinical observation of seizures. Green (2021) recommended nurses adopt a standardised approach to recognition of post-stroke complications, including recognition of seizures, but does not provide details on the approach to use (19). Bautista (2020) recommended a systematic assessment of seizures once recognised and provides details on key assessment areas: level of consciousness, eye deviation, gaze, pupil size, urinary incontinence, body movements and motor function (24). The onset and duration of seizure is recommended to be recorded along with neurological assessment continuously for the first five minutes of the ictal phase, and subsequent periodic assessments in ictal and postictal phases until the patient has returned to their baseline.
iv. Video recording. Mader’s (2020) case report describes video recording to seizure observation and diagnosis (22). The patient was video recorded alongside cEEG. The paper reports on a 28 second clonic seizure observed on video but, due to movement artefact, unable to be identified on EEG. This case also draws attention to the narrow time window to observe seizures if relying on human observation.
v. Family witness. Mader’s (2020) case report also highlights the contribution of relatives in observing seizure activity (22). The patient’s husband noticed her first post-stroke seizure - a 30 second episode of bilateral leg jerking 30 minutes after suspected brain stem stroke. This was the only paper referring to family’s contribution to recognition of seizure.

## DISCUSSION

To our knowledge, this is the first scoping review on the clinical methods used with acute stroke patients to identify and observe EPSS. We included eight papers: two research papers, two clinical guidelines and four discussion papers. We found no evaluation of different methods aimed at recognising and observing EPSS, and subsequently recommendations lacking detail and consensus on clinical processes. Continuous EEG is the method most referred to in the literature with detailed information of how to conduct observations at the bedside from critical care papers. Capturing seizure activity, either witnessed directly or by video recording, is key for contributing to accurate diagnosis and evaluating treatment. This is challenging when seizure activity can be subtle and only for a few seconds. This review identified that trained clinician observation, video recordings and relative witnesses all may have a role. More research into effective methods to capture observed seizure activity on acute stroke units, including support from relatives and the stroke multidisciplinary team, may be of value in improving seizure recognition.

We found in screening papers for the larger review, most available literature is concerned with the epidemiology and management of EPSS, with an assumption that post-stroke seizures are recognised by clinicians to start diagnostic investigations. More studies using EEG investigating seizure prevalence and characteristics in acute stroke were retrieved in our searches, but these papers were excluded as they did not provide detail on the assessment procedure, nor what methods were used to identify patients with suspected seizure activity. Whilst there is literature using clinical recognition and observation methods, such as EEG and clinical observations, our review has highlighted a lack of attention, particularly in the nursing literature, on the most effective and accurate methods for acute stroke patients.

We conducted a scoping review rather than a systematic review due to the lack of consensus in the literature on definition of EPSS. We aimed to include a wide range of literature using a systematic search process in extensive databases and within grey literature, but it is possible that we have missed some relevant literature. We did not undertake a formal quality assessment, but we did chart data on methodological information that informed our interpretation of the evidence. We did exclude papers that had mixed early and late seizure onset participants or where onset of seizure after stroke was not clear.

## Conclusions

Research on the prevalence, diagnosis and management of EPSS relies on effective recognition and observation of post-stroke patients for seizures. A lack of evidence on methods supporting identification and observation of seizures in post-stroke patients may contribute to underestimation of its prevalence and result in delayed diagnosis, increased complications and mortality. There is a need for more attention in research and clinical practice into consistent, systematic observation for EPSS and which methods, or combination of methods, might improve recognition rates of suspected seizure activity and ultimately improved diagnosis.

## Data Availability

All data produced in the present study are available upon reasonable request to the authors.

## Appendix 1: Preferred Reporting Items for Systematic reviews and Meta-Analyses extension for Scoping Reviews (PRISMA-ScR) Checklist

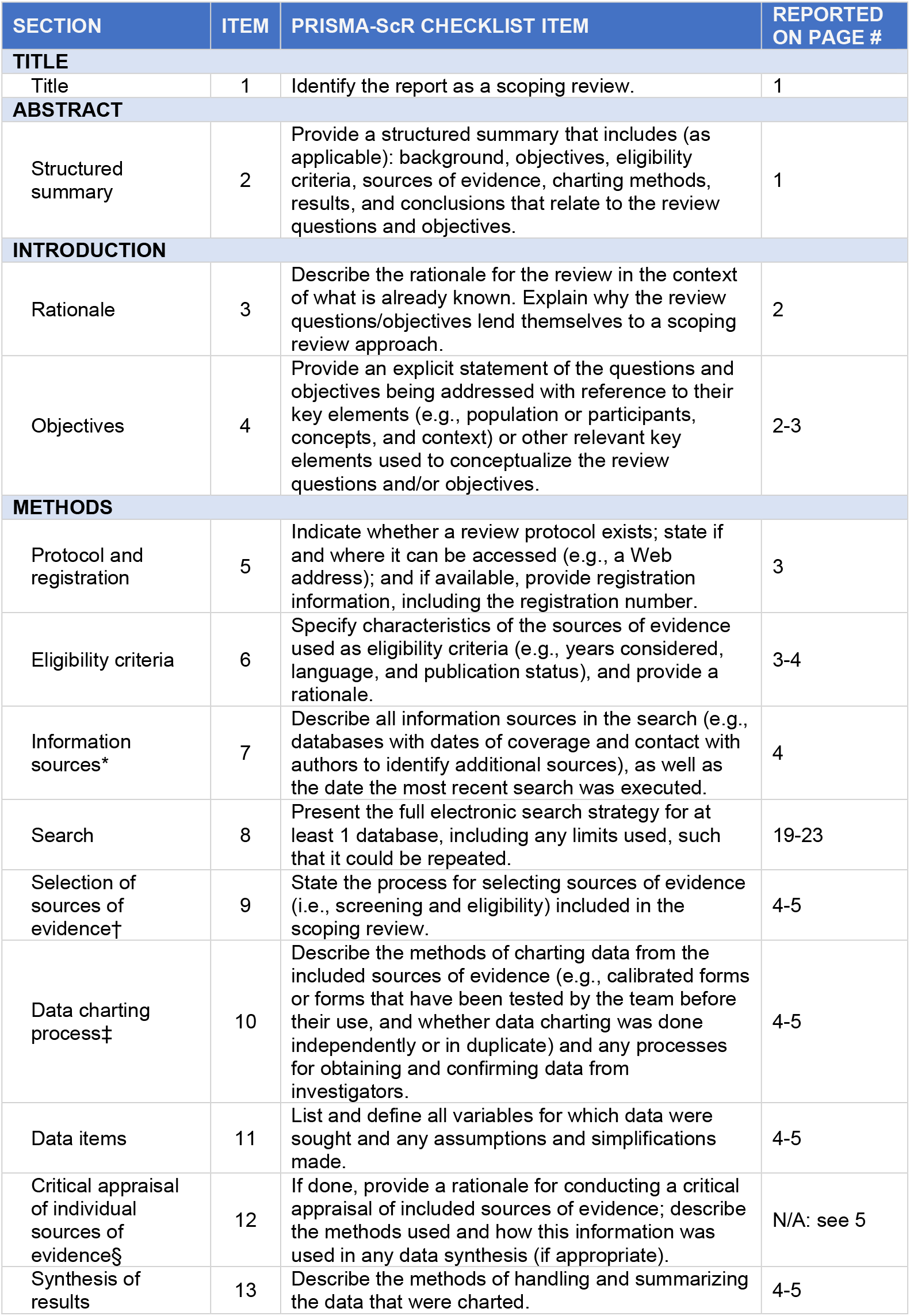
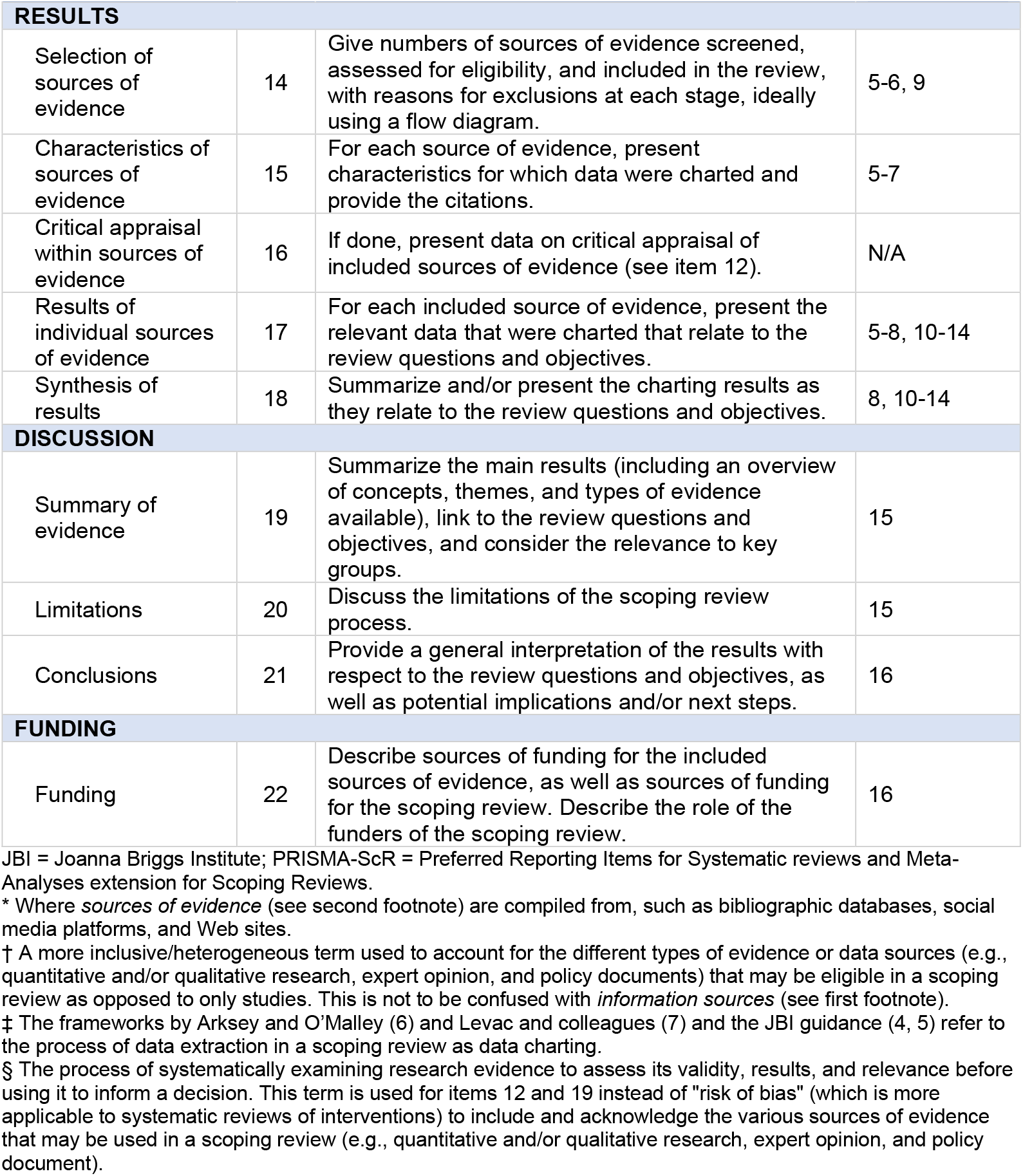

*From:* Tricco AC, Lillie E, Zarin W, O’Brien KK, Colquhoun H, Levac D, et al. PRISMA Extension for Scoping Reviews (PRISMAScR): Checklist and Explanation. Ann Intern Med. 2018;169:467–473. doi: 10.7326/M18-0850.

## Appendix 2: Search Strategies

### Database

**Ovid MEDLINE(R) and Epub Ahead of Print, In-Process, In-Data-Review & Other Non-Indexed Citations and Daily 1946 to September 30, 2021**

Date searched: 01/10/2021

1. Epilepsy/
2. Seizures/
3. Status Epilepticus/
4. 1 or 2 or 3
5. exp Stroke/
6. exp Cerebral Hemorrhage/
7. 5 or 6
8. 4 and 7
9. (Post-stroke seizure* or Poststroke seizure* or Post-stroke epilep* or Poststroke epilep* or postisch?emic stroke epilep* or post-isch?emic stroke epilep* or postisch?emic stroke seizure* or post-isch?emic stroke seizure* or postisch?emic seizure* or post-isch?emic seizure* or postisch?emic epilep* or post-isch?emic epilep* or posth?emorrhagic stroke epilep* or post-h?emorrhagic stroke epilep* or posth?emorrhagic stroke seizure* or post-h?emorrhagic stroke seizure* or acute symptomatic seizure*).ti,ab,kw.
10. ((seizure* or convuls* or epilep*) and (stroke* or poststroke or post-stroke or cerebrovascular accident* or cva or intracerebral h?emorrhage* or cerebral h?emorrhage* or brain h?emorrhage* or cerebral infarction* or brain infarction*)).ti.
11. ((seizure* or epilep* or convuls*) adj6 (stroke* or poststroke or post-stroke or cerebrovascular accident* or cva or intracerebral h?emorrhage* or cerebral h?emorrhage* or brain h?emorrhage* or cerebral infarction* or brain infarction*) adj6 (related or associated or follow* or after or onset or caus* or during or occur* or within)).ab.
12. 8 or 9 or 10 or 11
13. limit 12 to english language
14. exp animals/ not humans.sh.
15. 13 not 14
16. (exp child/ or exp infant/ or exp adolescent/) not exp Adult/
17. 15 not 16

### Database: Embase (Ovid) 1974 to 2021 September 30

Date searched: 01/10/2021

1. epilepsy/
2. seizure/
3. epileptic state/
4. 1 or 2 or 3
5. cerebrovascular accident/
6. brain infarction/
7. brain hemorrhage/
8. 5 or 6 or 7
9. 4 and 8
10. (Post-stroke seizure* or Poststroke seizure* or Post-stroke epilep* or Poststroke epilep* or postisch?emic stroke epilep* or post-isch?emic stroke epilep* or postisch?emic stroke seizure* or post-isch?emic stroke seizure* or postisch?emic seizure* or post-isch?emic seizure* or postisch?emic epilep* or post-isch?emic epilep* or posth?emorrhagic stroke epilep* or post-h?emorrhagic stroke epilep* or posth?emorrhagic stroke seizure* or post-h?emorrhagic stroke seizure* or acute symptomatic seizure*).ti,ab,kw.
11. ((seizure* or convuls* or epilep*) and (stroke* or poststroke or post-stroke or cerebrovascular accident* or cva or intracerebral h?emorrhage* or cerebral h?emorrhage* or brain h?emorrhage* or cerebral infarction* or brain infarction*)).ti.
12. ((seizure* or epilep* or convuls*) adj6 (stroke* or poststroke or post-stroke or cerebrovascular accident* or cva or intracerebral h?emorrhage* or cerebral h?emorrhage* or brain h?emorrhage* or cerebral infarction* or brain infarction*) adj6 (related or associated or follow* or after or onset or caus* or during or occur* or within)).ab.
13. 9 or 10 or 11 or 12
14. limit 13 to english language
15. (rat or rats or mouse or mice or swine or porcine or murine or sheep or lambs or pigs or piglets or rabbit or rabbits or cat or cats or dog or dogs or cattle or bovine or monkey or monkeys or trout or marmoset$1).ti. and animal experiment/
16. Animal experiment/ not (human experiment/ or human/)
17. 15 or 16
18. 14 not 17
19. (exp child/ or exp adolescence/ or exp adolescent/) not exp adult/
20. 18 not 19

### Database: CINAHL Complete (via EBSCOhost)

Date searched: 01/10/2021

S1 (MH “Epilepsy”)
S2 (MH “Seizures”)
S3 (MH “Status Epilepticus”) S4 S1 OR S2 OR S3
S5 (MH “Stroke+”)
S6 (MH “Cerebral Hemorrhage+”) S7 S5 OR S6
S8 S4 AND S7
S9 “Post-stroke seizure*” or “Poststroke seizure*” or “Post-stroke epilep*” or “Poststroke epilep*” or “postisch#emic stroke epilep*” or “post-isch#emic stroke epilep*” or “postisch#emic stroke seizure*” or “post-isch#emic stroke seizure*” or “postisch#emic seizure*” or “post-isch#emic seizure*” or “postisch#emic epilep*” or “post-isch#emic epilep*” or “posth#emorrhagic stroke epilep*” or “post-h#emorrhagic stroke epilep*” or “posth#emorrhagic stroke seizure*” or “post-h#emorrhagic stroke seizure*” or “acute symptomatic seizure*”
S10 TI ((seizure* or convuls* or epilep*) and (stroke* or poststroke or “post-stroke” or “cerebrovascular accident*” or cva or “intracerebral h#emorrhage*” or “cerebral h#emorrhage*” or “brain h#emorrhage*” or “cerebral infarction*” or “brain infarction*”))
S11 AB ((seizure* or epilep* or convuls*) N6 (stroke* or poststroke or “post-stroke” or “cerebrovascular accident*” or cva or “intracerebral h#emorrhage*” or “cerebral h#emorrhage*” or “brain h#emorrhage*” or “cerebral infarction*” or “brain infarction*”) N6 (related or associated or follow* or after or onset or caus* or during or occur* or within))
S12 S8 OR S9 OR S10 OR S11
S13 MH animals+
S14 MH (animal studies)
S15 TI (animal model*)
S16 S13 OR S14 OR S15
S17 MH (human)
S18 S16 NOT S17
S19 S12 NOT S18
S20 (MH “Child+”)
S21 (MH “Infant+”)
S22 (MH “Adolescence+”)
S23 S20 OR S21 OR S22
S24 (MH “Adult+”)
S25 S23 NOT S24
S26 S19 NOT S25
S27 S19 NOT S25 limited to English Language

### Database: Cochrane Library via Wiley (all databases)

Date searched: 01/10/2021

#1 MeSH descriptor: [Epilepsy] explode all trees
#2 MeSH descriptor: [Seizures] this term only
#3 MeSH descriptor: [Status Epilepticus] this term only
#4 #1 OR #2 OR #3
#5 MeSH descriptor: [Stroke] explode all trees
#6 MeSH descriptor: [Cerebral Hemorrhage] explode all trees #7 #5 OR #6
#8 #4 AND #7
#9 ((“post stroke” or poststroke or postischemic or postischaemic or “post-ischemic” or “post-ischaemic” or “postischemic Stroke” or “post-ischemic Stroke” or “postischaemic stroke” or “post-ischaemic stroke” or “posthemorrhagic stroke” or “post-hemorrhagic stroke” or “posthaemorrhagic stroke” or “post-haemorrhagic stroke”) NEXT (epilep* or seizure*)):ti,ab,kw
#10 (“acute symptomatic seizure” or “acute symptomatic seizures”):ti,ab,kw
#11 ((seizure* or convuls* or epilep*) and (stroke* or poststroke or “post-stroke” or (cerebrovascular NEXT accident*) or cva or (intracerebral NEXT h?emorrhage*) or (cerebral NEXT h?emorrhage*) or (brain NEXT h?emorrhage*) or (cerebral NEXT infarction*) or (brain NEXT infarction*))):ti
#12 (((seizure* or epilep* or convuls*) NEAR/6 (stroke* or poststroke or “post-stroke” or (cerebrovascular NEXT accident*) or cva or (intracerebral NEXT h?emorrhage*) or (cerebral NEXT h?emorrhage*) or (brain NEXT h?emorrhage*) or (cerebral NEXT infarction*) or (brain NEXT infarction*)) NEAR/6 (related or associated or follow* or after or onset or caus* or during or occur* or within))):ab
#13 #8 OR #9 OR #10 OR #11 OR #12
#14 MeSH descriptor: [Animals] explode all trees
#15 MeSH descriptor: [Humans] explode all trees
#16 #14 NOT #15
#17 #13 NOT #16
#18 MeSH descriptor: [Child] explode all trees
#19 MeSH descriptor: [Infant] explode all trees
#20 MeSH descriptor: [Adolescent] explode all trees
#21 #18 OR #19 OR #20
#22 MeSH descriptor: [Adult] explode all trees
#23 #21 NOT #22
#24 #17 NOT #23

